# Evaluation of clinical scales among populations diagnosed with atopic dermatitis: A scoping review

**DOI:** 10.1101/2025.09.02.25334962

**Authors:** Joy Xu, Sahrish Masood, Harshdeep Dhaliwal, Adil Amarsi, Amber Nelson-Fuller, Alexis Hoang, Hamza Naqvi, Maya Barua, Alyssa Wu, Sharon E. Albers

## Abstract

**Background:** Common diagnostic tools for atopic dermatitis (AD) often perform worse in skin-of-colour (SOC) populations. The objective of this review is to map the prevalence, validation, and effectiveness of clinician-based and patient-reported tools for diagnosing AD in SOC groups across all ages.

**Methods:** This review followed PRISMA-ScR guidelines and searched Embase, Scopus, PubMed, MEDLINE, Web of Science, and MedRxiv for articles published January 2015 through December 2023. Eligible studies were observational, randomized, or review articles evaluating clinician-rated scales or patient-reported measures with self-identified race or ethnicity. We excluded non-English publications, case reports/series, guidelines, editorials, and studies lacking stratification. After de-duplication, two reviewers screened titles, abstracts, and full texts with conflicts resolved by a third reviewer. Data extraction captured study design, population demographics, tools evaluated, and key findings on accuracy and reliability in SOC cohorts.

**Results:** 28 articles (total n = 20 332) met inclusion criteria. 24 assessed clinician-rated scales, most often EASI (n = 16), SCORAD (n = 10), and o-SCORAD (n = 8). These tools frequently underestimate AD severity in Fitzpatrick IV-VI skin types. Five studies examined alternative clinician tools (vIGA-AD, IGAxBSA). Rajka-Langeland and ADSI scores were each assessed once. Patient-reported outcomes (PROs) were dominated by POEM (n = 17), which had only 14% SOC participants during initial validation. PO-SCORAD (a PRO based on SCORAD) was also assessed (n = 10). Nine newer PRO tools (RECAP, ADCT, PSAAD, ADSEQ, CEQ, DFI, CADIS, QoLIAD, PIQoL-AD) appeared in single studies. Adjunctive measures and technological approaches (body-surface area alone, photo guides, AI-assisted analysis, remote assessment) featured in five studies but lack multi-center validation.

**Conclusions:** Most diagnostic tools remain validated in lighter-skinned cohorts and underrepresent SOC populations. Patient-reported measures show promise but require wider validation. Adjunctive and technology-driven methods may improve equity but need rigorous testing. Future research should prioritize multiethnic cohorts, age-specific validation, and consensus-driven adaptation of both clinician and patient-reported tools to ensure reliable assessment for all skin types.

## Introduction

Atopic dermatitis (AD) is a chronic inflammatory skin disease that often precedes other allergic conditions such as asthma or allergic rhinitis ^1^. The etiology of AD arises from a combination of genetic predispositions, including mutations impairing the skin barrier, immune dysregulation, and environmental factors ^1–3^. Indications include chronic or recurrent pruritus, erythema, and in many cases, secondary infections. Acute flare-ups vary by individual with an array of triggers including stress, climate, sweating, and allergens ^4^.

AD afflicts approximately 245 million individuals worldwide, with a pediatric prevalence of 20.1% and adult prevalence of 9.7% ^5^. This renders it the most common form of eczema ^1,6^. In the United States, Black and Hispanic children in the United States are disproportionately affected by atopic dermatitis, experiencing higher prevalence, greater disease severity, earlier onset, and more persistent disease compared with White children ^7^. These disparities are shaped primarily by environmental exposures, healthcare access, and structural racism rather than genetic differences. Moreover, Black and Hispanic patients experience severe, persistent disease, and even report 1.8- to 2.3-fold higher annual healthcare expenditures compared to their White counterparts ^8–13^. These patients also report diminished quality of life due to limitations imposed on daily activities in domestic and occupational settings, with additional risk of hospitalization and higher health care expenditures ^14,15^. This substantial epidemiologic and economic burden underscores the imperative for accurate and validated assessment tools that perform reliably across all demographic groups, including skin of color (SOC) populations.

Clinician-rated scales, notably Eczema Area and Severity Index (EASI) and Scoring Atopic Dermatitis (SCORAD), are most frequently used to diagnose AD. However, their performance across demographics, especially those with SOC, remains limited. In the LEAD validation trial for EASI (n = 500), only 38% of participants were Fitzpatrick IV–VI, and inter-rater reliability in these types averaged a 1.2 point underestimation compared to Fitzpatrick I–III ^16^. SCORAD’s sensitivity also falls sharply, from 82% in I–III to 64% in IV–VI (p<0.01) ^17^. Thus, reliability of these tools greatly decreases with increased skin pigmentation, highlighting the urgent need for better validation or assessment tools.

Patient- and parent-reported outcome measures, such as the Patient Oriented Eczema Measure (POEM), can capture symptom fluctuations missed by clinician scales. However, to date, POEM has only been validated in cohorts with limited SOC participants; for example, only 14% patients in the original development study by Charman et al. had SOC. Alternative approaches include Artificial Intelligence (AI)-driven image-analysis algorithms, showing some promise to address existing gaps ^18^. Over 15 devices are regulatory-approved worldwide (including three FDA-cleared systems in the U.S.), and early studies suggest these algorithms can standardize severity scoring across diverse skin tones ^19^.

Despite these advances, all current tools rely heavily on visually-intensive assessment of erythema and lichenification presentation. These features are confounded with distinct presentations across varying levels of pigmented skin, and can present in lesser-known ways in patients with SOC ^20,21^. As most tools were validated in lighter skinned cohorts, evidence for their reliability in diverse populations remains limited ^22^.

Given these prevailing disparities, the objective of this scoping review is to systematically assess the availability, validation, and effectiveness of both clinician-based and patient or caregiver reported diagnostic tools for AD in SOC populations across age groups.

## Methods

This review was originally registered in 2024 with PROSPERO (CRD42024553332) as a systematic review for scales diagnosing skin cancer, but was later reestablished and reconducted as a scoping review for scales diagnosing AD. This decision was made given a paucity of race/ethnicity-stratified validation data and the strong relevance of this topic for AD. This review was conducted according to PRISMA-ScR guidelines. Inclusion criteria were: (1) observational, randomized, or review articles published January 2015–December 2023; (2) evaluation of clinician-based or patient-reported AD diagnostic tools; (3) race/ethnicity measures; and (4) data from the U.S. and/or Canada. Exclusion criteria were: non-English, case reports/series, guidelines, editorials, and lack of stratification. The Embase, Scopus, PubMed, MEDLINE, Web of Science, and MedRxiv databases were searched with strings such as [(“atopic derm*”) AND (“diagnos*”) AND (“quality of life”) AND ((“tool”) OR (“scale”) OR (“assess*)) NOT ((“artificial intelligence”) OR (“ai”))].

Hand searching was primarily conducted through an automated web scraping python script. The script looked for included articles’ PubMed listings; both the similar articles section and reference list were scraped for articles. Any with a PubMed link were followed and saved for screening. The code used for automation is available upon request. ResearchRabbit, a citation-based literature mapping AI tool, was also used to supplement the hand search. See figure 1 for the PRISMA flowchart for this review. After de-duplication, two reviewers independently screened titles/abstracts and full texts, with disagreements resolved by a third reviewer through Covidence. Data extraction was performed using a pre-piloted collection form standardized to all reviewers. Extracted data included study design, methodology, year, location, sample size, population, demographics, diagnostic tools evaluated (clinician-based and PROs), phenotypic characteristics (such as Fitzpatrick skin type or ethnicity), and key findings regarding diagnostic accuracy, reliability, and usability in SOC populations.

**Figure 1.**
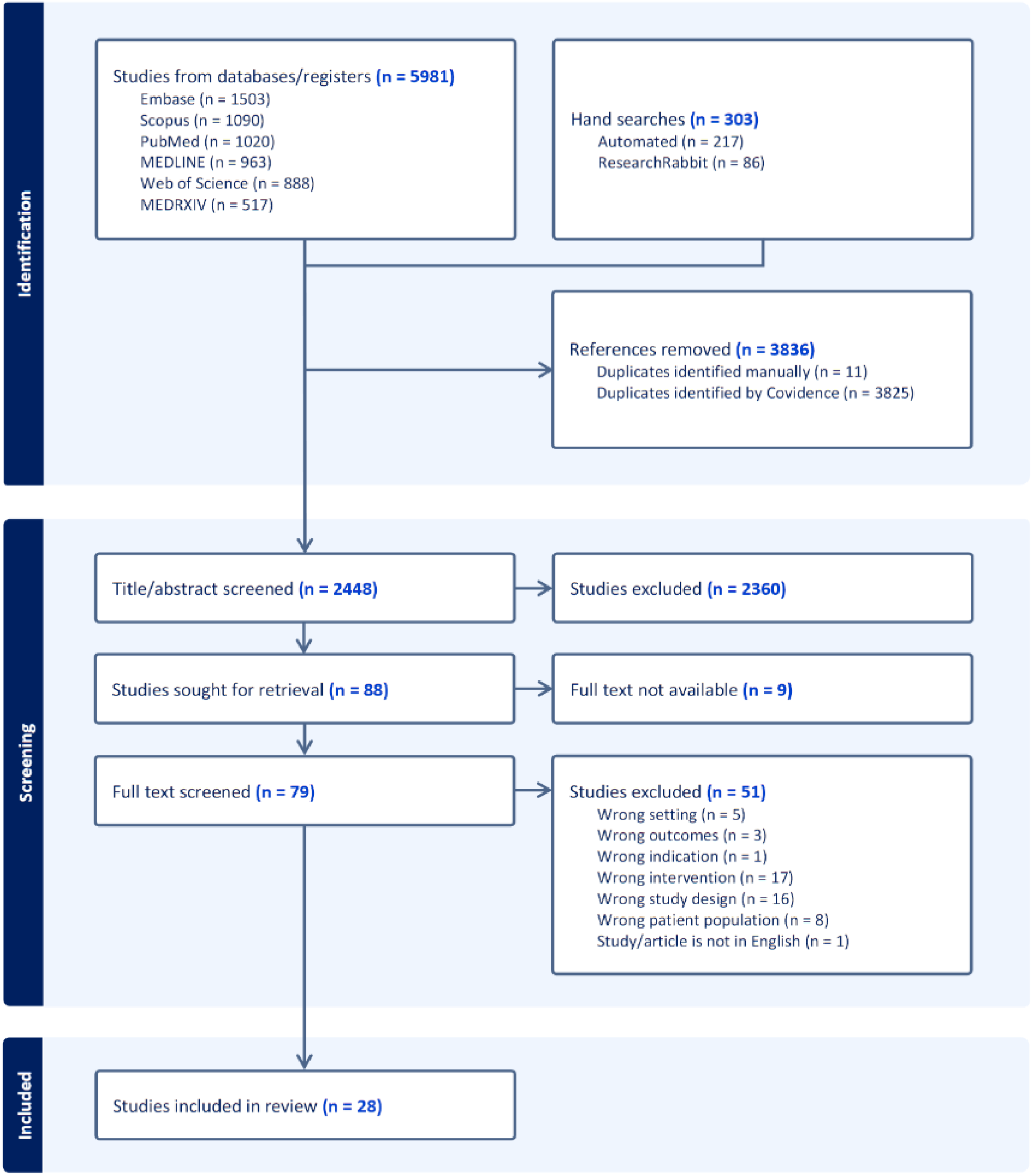
PRISMA Flowchart

## Results

### Overall Landscape

28 studies and reviews (total n = 20 332) met inclusion criteria. Most studies (n = 27) were U.S.-based. Study designs consisted of prospective cohort (n = 13), cross-sectional (n = 7), retrospective study (n = 2), validation (n = 3), literature review (n = 2), and post-hoc analysis (n = 1). Table 1 gives an overview of each article’s metadata, patient demographics, and AD-specific scales assessed. Given the heterogeneity of populations, methods, and outcome measures, meta-analysis was precluded. Findings were synthesized narratively, focusing on each tool’s diagnostic strengths, limitations, and validation gaps in SOC cohorts.

Across 28 studies of atopic dermatitis severity tools, clinician-rated scales appeared in 24 studies (83%), patient-reported outcomes appeared in 16 studies (55%), and adjunctive or technological measures appeared in five studies (17%). Table 2 details scoring methods for each scale. Figure 2 summarizes the occurrences of each scale in extracted studies. The overall emphasis on clinician tools indicates a reliance on observed patient signs. However, patient reported outcomes which also assess patient symptoms are growing and are included in over half of the assessed recent literature. Newer methods such as imaging guides and artificial intelligence remain limited.

**Figure 2.**
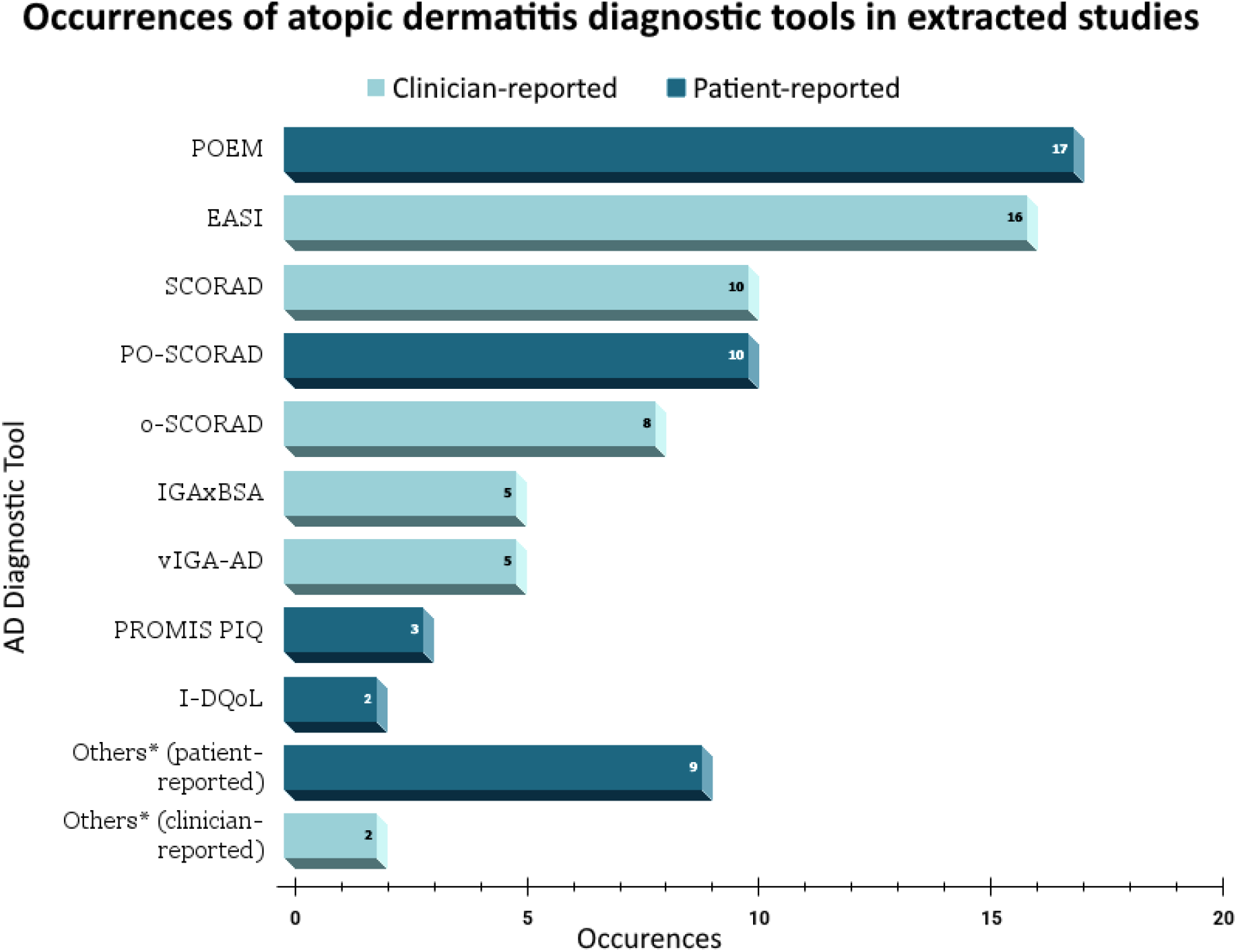
Occurrences of atopic dermatitis diagnostic tools in extracted studies *The ‘Others’ category represents tools mentioned only once in the extracted studies. The other clinician-reported tools consist of the Rajka–Langeland severity score and ADSI (Atopic Dermatitis Severity Index). The other patient-reported tools consist of the RECAP (Recap of Atopic Eczema); ADCT (Atopic Dermatitis Control Tool); PSAAD (Pruritus and Symptoms Assessment for Atopic Dermatitis); ADSEQ (Atopic Dermatitis Screening and Evaluation Questionnaire); CEQ (Childhood Eczema Questionnaire); DFI (Dermatitis Family Impact Questionnaire); CADIS (Childhood Atopic Dermatitis Impact Scale); QoLIAD (Quality of Life Index for Atopic Dermatitis); and PIQoL-AD (Parents’ Index of Quality of Life in Atopic Dermatitis).

### Clinician-rated scales

The Eczema Area and Severity Index (EASI) was used in 16 studies. Each body region is graded 0 to 3 for redness, thickness, scratching changes and skin texture, then weighted and summed to yield 0 to 72. Higher scores indicate more severe and widespread disease. Scoring Atopic Dermatitis (SCORAD) and objective SCORAD (o-SCORAD) appeared in 10 and 8 studies respectively. SCORAD combines percent of body-surface area with six signs scored 0 to 3 and two patient symptom ratings scored 0 to 10 to yield 0 to 103. o-SCORAD excludes subjective components. PO-SCORAD, a PRO which is a validated patient self-assessment modification of SCORAD, appeared in 10 studies. PO-SCORAD shows a strong correlation with SCORAD. Both EASI and SCORAD reproduced clinician impressions but undercounted severity in darker skin types. Only a minority of EASI and SCORAD studies (fewer than 20%) reported how many participants had Fitzpatrick skin types IV to VI, pointing to a critical gap in scale inclusivity.

### Alternative clinician tools

The Validated Investigator Global Assessment for Atopic Dermatitis (vIGA-AD) was evaluated in five studies. Clinicians assign a single score from 0 (clear) to 4 (severe). This simplicity can speed ratings but may mask subtle changes in diverse skin tones. The Investigator Global Assessment multiplied by percent body surface area (IGA×BSA) also appeared in five studies. It multiplies a 0 to 4 global grade by the percent of skin affected for a 0 to 400 range. This composite captures both extent and intensity but was rarely tested in participants with darker skin. Two studies of the Rajka-Langeland score and two of the Atopic Dermatitis Severity Index each enrolled fewer than 700 patients. These brief multi-item indices remain unvalidated in underrepresented groups.

### Patient-reported outcomes (PROs)

The Patient-Oriented Eczema Measure (POEM) was assessed in 17 studies. It asks how often seven symptoms occurred over the last week, scoring each from 0 (never) to 4 (every day) for a total of 0 to 28. Lower POEM scores can miss rare but severe flares, while higher scores map to consistent symptom burden. Only 14% of the roughly 1 158 POEM participants in the initial validation study were those with SOC, suggesting that symptom experiences in more pigmented populations are underreported. Other frequently reported tools in the included studies, such as the Dermatology Life Quality Index and Skindex-29, are not AD specific and assess health-related quality of life. Additionally, several AD PROs in the included literature were mentioned only once: Recap of Atopic Eczema; Atopic Dermatitis Control Tool; Pruritus and Symptoms Assessment for Atopic Dermatitis; Atopic Dermatitis Screening and Evaluation Questionnaire; Childhood Eczema Questionnaire; Dermatitis Family Impact Questionnaire; Childhood Atopic Dermatitis Impact Scale; Quality of Life Index for Atopic Dermatitis; and Parents’ Index of Quality of Life in Atopic Dermatitis. Their single appearances may indicate growing interest in improving patient-reported measurement.

### Adjunctive measures

Body-surface area alone was used in three studies to quantify extent but never validated as a stand-alone severity marker. Two studies tested photo reference guides for darker skin tones and found improved clinician detection of subtle redness, but no universal atlas exists. AI-assisted image analysis and remote assessment via caregiver photos or videos appeared in three studies. Preliminary algorithms show promise at standardizing severity scores across skin types. However, none of these methods have completed multi-center validation with balanced representation of Fitzpatrick types I through VI.

### Technological innovations

A small number of studies described AI-assisted image analysis and remote assessment methods. Early reports suggest these approaches may standardize severity scoring across skin tones, but comprehensive validation in diverse cohorts is pending.

### Interpretation

The clinician-rated scales EASI and SCORAD remain the backbone of severity assessment but often overlook subtleties in pigmented skin. Less than one in five studies explicitly tracked Fitzpatrick IV to VI participants, which may indicate a strong bias in the AD literature toward lighter skin. Patient-reported measures such as POEM capture symptoms that clinicians miss, yet underrepresent diverse populations. Adjunctive and technological tools have potential to close these gaps but require focused validation in inclusive cohorts.

## Discussion

This scoping review highlights persistent diagnostic inequities in AD, especially for individuals with SOC. EASI and SCORAD, the most widely used clinician-based diagnostic scales, remain fundamentally limited by their reliance on visual assessment of erythema and lichenification. These features can be less distinct or may appear differently in darker skin tones, sometimes presenting as violaceous brown or subtle color changes that are easily missed by clinicians who have been trained mainly on lighter skin ^20,23^. Several studies included in this review reported that erythema is not only more difficult to detect in Fitzpatrick types IV to VI but that lichenification and post-inflammatory pigmentary changes often dominate the presentation of AD in SOC. Pigmentary changes such as hyperpigmentation and hypopigmentation are not currently captured by most existing scoring systems ^20,23,24^. Consequently, clinicians may underestimate disease severity which can lead to undertreatment and a reduced likelihood of appropriate referral or escalation of care for patients with SOC.

Few diagnostic tools have been robustly validated in ethnically diverse cohorts ^20^. Some studies have proposed adapting scoring systems to account for post-inflammatory hyperpigmentation periorbital dark circles and flexural accentuation, features more common or pronounced in SOC ^23,24^. However, these adaptations are inconsistently applied and have not been standardized in routine clinical practice ^20^. Unique presentations such as lichen planus-like AD and periorbital darkening occur more frequently in SOC but are rarely addressed in validation studies ^23^. This lack of inclusive validation continues to perpetuate disparities in diagnosis and disease monitoring.

In parallel, PRO measures such as POEM have received increasing attention as complementary tools for assessing disease burden. These measures are especially valuable in cases where objective clinical signs are ambiguous or difficult to interpret, which is often the situation for patients with SOC ^12^. This review found that most PROs, including POEM, have not been systematically validated across diverse skin types or languages ^15^. As a result, questions remain about whether these instruments fully capture the specific symptoms, burdens and impact experienced by SOC populations. For example, pruritus dyspigmentation or burning sensations may be more prominent in SOC and could be underemphasized in standard questionnaires ^15^.

The need for robust and inclusive tools is especially acute in pediatric populations. Younger children are often unable to self-report symptoms reliably, making parent or caregiver-reported tools critical for accurate assessment. However, few studies have evaluated these measures specifically in SOC children or across different cultural backgrounds ^10^. Pediatric AD in SOC can also present differently compared to adults and may be mistaken for other dermatoses further compounding the risk of misdiagnosis or underestimation of severity ^11^. Developmental differences in symptom perception and expression as well as the involvement of caregivers in reporting highlight the need for further adaptation and validation of existing pediatric tools to ensure equity.

Technological advances offer additional opportunities and challenges. Some studies suggested that using BSA assessment and real-life photo references may improve diagnostic accuracy for SOC, but systematic implementation and validation of these strategies remain limited. Innovations in artificial intelligence and remote teledermatology show promise for reducing diagnostic disparities. With current AI algorithms trained primarily on images of lighter skin, this limits their accuracy and generalizability when applied to patients with SOC ^25^.Only one study in this review specifically evaluated AI-based tools in a diverse skin color cohort and few telehealth platforms have been examined for diagnostic equity. The lack of standardized and diverse photo reference sets remains a barrier to widespread adoption.

Broader trends in the literature reveal that clinician-based tools like EASI and SCORAD have dominated the diagnostic landscape despite their limited applicability for SOC populations. While newer instruments such as vIGA-AD and IGAxBSA are being explored, the lack of validation and standardization for SOC remains a significant obstacle ^22^. PROs show promise for improving equity, but further development is necessary to ensure these tools can detect the full spectrum of disease presentations in diverse populations. There is an ongoing need for adaptation of existing tools and the creation of new metrics that reflect both lived experience and phenotypic variability in SOC.

Efforts to diversify research cohorts and include underrepresented populations are essential for future progress. Strategies such as targeted recruitment, community partnerships, and culturally sensitive research design should be prioritized to make research findings more generalizable. SOC-inclusive photo references and expanded validation of both clinician and patient-reported tools should be at the forefront of research agendas. Investments in scalable and cost-effective solutions including digital tools, AI-driven models with diverse training sets, and remote assessment platforms are crucial for improving care in resource-limited settings where diagnostic inequity is most pronounced.

## Conclusion

Current diagnostic tools for AD remain insufficient for accurate and equitable assessment in SOC populations, largely because of the continued reliance on clinician-based evaluation of erythema and the lack of scale adaptation for diverse skin types. Broader use of patient- and caregiver-reported outcomes, incorporation of phenotypic variability, and integration of emerging technological solutions have the potential to advance equity. However, these approaches must undergo robust validation and standardization. In summary, this review underscores that diagnostic tools for AD have not kept pace with the diversity of patient populations. Achieving equity in AD diagnosis will require collaborative efforts among clinicians, researchers, and communities to validate, adapt, and widely implement inclusive and reliable assessment tools. Future research should prioritize multiethnic cohorts, age-specific measures, and consensus-driven adaptations to ensure that diagnostic innovation benefits all individuals with AD.

## Supporting information

Table 1: Included articles summary

Table 2: Atopic dermatitis scales overview

## Data Availability

All data produced in the present study are available upon reasonable request to the authors. Key data is present in Table 1 and 2.

