## Supplementary material for "Evaluation of clinical scales among populations diagnosed with atopic dermatitis: A scoping review": Table 1: Included articles summary

| Article Information | | | | | Patient Demographics | | | Scales |
|---|---|---|---|---|---|---|---|---|
| Study Name | Study Design | First Author | Year | Country | Sample Size | Age, Mean (SD) | Sex (Female %) | AD Diagnostic/Severity Scales Included |
| Comparison of Patient-Oriented Eczema Measure and Patient-Oriented Scoring Atopic Dermatitis vs Eczema Area and Severity Index and other measures of atopic dermatitis: A validation study. | Prospective Cohort Study | Silverberg, JI | 2020 | USA, Switzerland | 291 | 39.9 (14.9) | 51.60% | PO-SCORAD (Patient-Oriented Scoring of Atopic Dermatitis)<br>POEM (Patient-Oriented Eczema Measure)<br>EASI (Eczema Area and Severity Index)<br>SCORAD (Scoring of Atopic Dermatitis) |
| Measuring Atopic Dermatitis Disease Severity: The Potential for Electronic Tools to Benefit Clinical Care. | Review (Literature) | Maintz, L | 2021 | Germany, Switzerland, Canada | N/A | N/A | N/A | EASI (Eczema Area and Severity Index)<br>POEM (Patient-Oriented Eczema Measure)<br>I-DQoL (Infant's Dermatitis Quality of Life Index)<br>RECAP (Recap of Atopic Eczema)<br>ADCT (Atopic Dermatitis Control Tool)<br>vIGA-AD (Validated Investigator Global Assessment for Atopic Dermatitis)<br>SCORAD (Scoring of Atopic Dermatitis)<br>o-SCORAD (objective SCORAD)<br>PO-SCORAD (Patient-Oriented Scoring of Atopic Dermatitis)<br>PSAAD (Pruritus and Symptoms Assessment for Atopic Dermatitis) |
| Product of Investigator Global Assessment and Body Surface Area (IGAxBSA): A practice-friendly alternative to the Eczema Area and Severity Index to assess atopic dermatitis severity in children. | Cross-Sectional Study | Suh, TP | 2020 | USA | 195 | 10.3 (3.5) | 58.50% | IGAxBSA (Product of Investigator Global Assessment and Body Surface Area)<br>EASI (Eczema Area and Severity Index)<br>vIGA-AD (Validated Investigator Global Assessment for Atopic Dermatitis)<br>o-SCORAD (objective SCORAD)<br>SCORAD (Scoring of Atopic Dermatitis)<br>POEM (Patient-Oriented Eczema Measure) |
| Severity strata for POEM, PO-SCORAD, and DLQI in US adults with atopic dermatitis. | Cross-Sectional Study | Silverberg, JI | 2018 | USA | 602 | 52.0 (16.3) | 53.60% | POEM (Patient-Oriented Eczema Measure)<br>PO-SCORAD (Patient-Oriented Scoring of Atopic Dermatitis) |
| Validation of five patient-reported outcomes for atopic dermatitis severity in adults. | Cross-Sectional Study | Silverberg, JI | 2019 | USA | 2893 | 52.0 (16.3) | 53.60% | PO-SCORAD (Patient-Oriented Scoring of Atopic Dermatitis)<br>POEM (Patient-Oriented Eczema Measure) |
| Validation of patient-reported global severity of atopic dermatitis in adults. | Prospective Cohort Study | Vakharia, PP | 2017 | USA | 265 | 42.5 (17.5) | 61.10% | o-SCORAD (objective SCORAD)<br>SCORAD (Scoring of Atopic Dermatitis)<br>EASI (Eczema Area and Severity Index)<br>POEM (Patient-Oriented Eczema Measure) |
| Validation of remote atopic dermatitis severity assessment with the Eczema Area and Severity Ondex in children using caregiver-provided photos and videos. | Cross-Sectional Study | Croce, EA | 2022 | USA | 50 | 4.3 (4.4) | 40.00% | EASI (Eczema Area and Severity Index) |
| IGAxBSA composite for assessing disease severity and response in patients with atopic dermatitis. | Post Hoc Analysis of RCTs and OLEs | Paller, AS | 2022 | USA, Canada | 3473 | 33.2 (16.4) [RCTs]; 35.5 (16.1) [OLEs] | RCTs: 42.50%; OLEs: 41.10% | IGAxBSA (Product of Investigator Global Assessment and Body Surface Area)<br>vIGA-AD (Validated Investigator Global Assessment for Atopic Dermatitis)<br>EASI (Eczema Area and Severity Index)<br>SCORAD (Scoring of Atopic Dermatitis)<br>o-SCORAD (objective SCORAD)<br>POEM (Patient-Oriented Eczema Measure) |

| Article Information | | | | | Patient Demographics | | | Scales |
|---|---|---|---|---|---|---|---|---|
| Study Name | Study Design | First Author | Year | Country | Sample Size | Age, Mean (SD) | Sex (Female %) | AD Diagnostic/Severity Scales Included |
| Measurement properties of the product of investigator's global assessment and body surface area in children and adults with atopic dermatitis. | Prospective Cohort Study | Silverberg, JI | 2020 | USA, Switzerland | 653 | 42.6 (19.3) | 56.20% | IGAxBSA (Product of Investigator Global Assessment and Body Surface Area)<br>EASI (Eczema Area and Severity Index)<br>o-SCORAD (objective SCORAD)<br>POEM (Patient-Oriented Eczema Measure)<br>vIGA-AD (Validated Investigator Global Assessment for Atopic Dermatitis) |
| Measurement properties of the Rajka-Langeland severity score in children and adults with atopic dermatitis. | Prospective Cohort Study | Silverberg, JI | 2020 | USA, Switzerland | 427 | 41.4 (17.9) | 57.40% | Rajka–Langeland severity score<br>EASI (Eczema Area and Severity Index)<br>SCORAD (Scoring of Atopic Dermatitis)<br>o-SCORAD (objective SCORAD)<br>POEM (Patient-Oriented Eczema Measure)<br>PIQ (PROMIS (Patient-Reported Outcomes Measurement Information System) Itch Questionnaire) |
| Pilot testing and validation of an atopic dermatitis screening and evaluation questionnaire. | Prospective Cohort Study | Chen, CA | 2016 | USA | 108 | NR | NR | ADSEQ (Atopic Dermatitis Screening and Evaluation Questionnaire) |
| Validation of a Parent-Reported Diagnostic Instrument in a U.S. Referral Population: The Childhood Eczema Questionnaire. | Validation Study | Leitenberger, S | 2017 | USA | 242 | NR | NR | CEQ (Childhood Eczema Questionnaire) |
| Construct validity of self-reported global atopic dermatitis severity in a population-based cohort of adults | Cross-sectional study | Silverberg, JI | 2018 | USA | 602 | 52.0 (16.3) | 53.60% | POEM (Patient-Oriented Eczema Measure)<br>PO-SCORAD (Patient-Oriented Scoring of Atopic Dermatitis) |
| Measurement properties of the Patient-Reported Outcomes Information System (PROMIS) Itch Questionnaire: itch severity assessments in adults with atopic dermatitis. | Validation Study | Silverberg, JI | 2020 | USA | 410 | 44.9 (18.3) | 63.20% | POEM (Patient-Oriented Eczema Measure)<br>PO-SCORAD (Patient-Oriented Scoring of Atopic Dermatitis)<br>EASI (Eczema Area and Severity Index)<br>PIQ (PROMIS (Patient-Reported Outcomes Measurement Information System) Itch Questionnaire) |
| Validity and reliability of Patient-Reported Outcomes Measurement Information System Global Health scale in adults with atopic dermatitis. | Prospective Cohort Study | Schwartzman, G | 2021 | USA | 994 | 44.7 (16.7) | 67.20% | PO-SCORAD (Patient-Oriented Scoring of Atopic Dermatitis)<br>EASI (Eczema Area and Severity Index)<br>POEM (Patient-Oriented Eczema Measure)<br>SCORAD (Scoring of Atopic Dermatitis) |
| What the Eczema Area and Severity Index score tells us about the severity of atopic dermatitis: an interpretability study | Retrospective Study | Leshem, YA | 2015 | USA | 170 | 15.2 (15.7) | 54.10% | EASI (Eczema Area and Severity Index) |
| Severity strata for Eczema Area and Severity Index (EASI), modified EASI, Scoring Atopic Dermatitis (SCORAD), objective SCORAD, Atopic Dermatitis Severity Index and body surface area in adolescents and adults with atopic dermatitis | Cross-sectional Study | Chopra, R | 2017 | USA | 673 | 43.4 (19.6) | 54.12% | EASI (Eczema Area and Severity Index)<br>IGAxBSA (Product of Investigator Global Assessment and Body Surface Area)<br>SCORAD (Scoring of Atopic Dermatitis)<br>ADSI (Atopic Dermatitis Severity Index) |

| Article Information | | | | | Patient Demographics | | | Scales |
|---|---|---|---|---|---|---|---|---|
| **Study Name** | **Study Design** | **First Author** | **Year** | **Country** | **Sample Size** | **Age, Mean (SD)** | **Sex (Female %)** | **AD Diagnostic/Severity Scales Included** |
| The Validated Investigator Global Assessment for Atopic Dermatitis (vIGA-AD): a clinical outcome measure for the severity of atopic dermatitis. | Validation Study | Simpson, EL | 2020 | USA | 1679 | 35.6 (12.8); 34.7 (12.8); 39.5 (16.1) [Three trials] | 37.30%; 38.00%; 49.10% [Three trials] | vIGA-AD (Validated Investigator Global Assessment for Atopic Dermatitis) |
| IGAxBSA composite for assessing disease severity and response in patients with atopic dermatitis. | Retrospective Study | Paller, AS | 2022 | USA | 3473 | 33.2 (16.6) | 42.50% | IGAxBSA (Product of Investigator Global Assessment and Body Surface Area)<br>o-SCORAD (objective SCORAD)<br>POEM (Patient-Oriented Eczema Measure)<br>EASI (Eczema Area and Severity Index)<br>SCORAD (Scoring of Atopic Dermatitis) |
| Content and construct validity, predictors, and distribution of self-reported atopic dermatitis severity in US adults. | Cross-sectional Study | Silverberg, JI | 2018 | USA | 602 | 52.0 (16.3) | 53.60% | PO-SCORAD (Patient-Oriented Scoring of Atopic Dermatitis)<br>POEM (Patient-Oriented Eczema Measure) |
| Patient-reported outcomes and quality of life measures in atopic dermatitis | Literature Review | Paras, P | 2018 | USA | N/A | N/A | N/A | DFI (Dermatitis Family Impact Questionnaire)<br>I-DQoL (Infant's Dermatitis Quality of Life Index)<br>CADIS (Childhood Atopic Dermatitis Impact Scale)<br>QoLIAD (Quality of Life Index for Atopic Dermatitis)<br>PIQoL-AD (Parents' Index of Quality of Life in Atopic Dermatitis)<br>PO-SCORAD (Patient-Oriented Scoring of Atopic Dermatitis) |
| Severity strata for five patient-reported outcomes in adults with atopic dermatitis | Prospective Cohort Study | Vakharia, PP | 2017 | USA | 210 | 39.8 (17.8) | 62.70% | POEM (Patient-Oriented Eczema Measure) |
| What are the best endpoints for Eczema Area and Severity Index and Scoring Atopic Dermatitis in clinical practice? A prospective observational study | Prospective Cohort Study | Silverberg, JI | 2019 | USA | 826 | 46.2 (19.3) | 52.50% | EASI (Eczema Area and Severity Index)<br>SCORAD (Scoring of Atopic Dermatitis)<br>o-SCORAD (objective SCORAD) |
| Validation of patient-reported global severity of atopic dermatitis in adults | Prospective Cohort Study | Vakharia, PP | 2017 | USA | 265 | 42.5 (17.5) | 61.10% | POEM (Patient-Oriented Eczema Measure)<br>PO-SCORAD (Patient-Oriented Scoring of Atopic Dermatitis)<br>EASI (Eczema Area and Severity Index) |
| Measurement properties of three assessments of burden used in atopic dermatitis in adults | Prospective Cohort Study | Patel, KR | 2019 | USA | 340 | 42.8 (16.5) | 67.40% | POEM (Patient-Oriented Eczema Measure) |
| Validation of remote atopic dermatitis severity assessment with the Eczema Area and Severity Ondex in children using caregiver provided photos and videos | Prospective Cohort Study | Croce, EA | 2023 | USA | 50 | 4.3 (4.4) | 42.00% | EASI (Eczema Area and Severity Index) |
| Development, Validation, and Interpretation of the PROMIS Itch Questionnaire: A Patient-Reported Outcome Measure for the Quality of Life Impact of Itch | Prospective Cohort Study | Silverberg, JI | 2020 | USA | 600 | NR | 62.00% | PIQ (PROMIS (Patient-Reported Outcomes Measurement Information System) Itch Questionnaire) |
| Measurement properties of the Patient-Reported Outcomes Measurement Information System Itch Questionnaire item banks in adults with atopic dermatitis. | Prospective Cohort Study | Silverberg, JI | 2020 | USA | 239 | 46.8 (18.2) | 62.30% | PIQ (PROMIS (Patient-Reported Outcomes Measurement Information System) Itch Questionnaire) |
