## Supplementary material for "Evaluation of clinical scales among populations diagnosed with atopic dermatitis: A scoping review": Table 2: Atopic dermatitis scales overview

| Name of Scale | Description | Age | Diagnoses | Scoring |
|---|---|---|---|---|
| PO-SCORAD (Patient-Oriented Scoring of Atopic Dermatitis) | Derived from SCORAD, PO-SCORAD is a patient-oriented self-assessment scale for patients with atopic dermatitis that scores severity of disease. PO-SCORAD permits continuous self-assessment by the patient, but it can also be used in clinical, hospital, and research settings. The criteria to determine use on patients include the diagnosis of atopic dermatitis based on Hanifin-Rajka criteria; and the patient's ability to self-assess and report the severity and extent of symptoms.It is structured into three parts, assessing the extent of disease, severity, and an evaluation of subjective symptoms. The extent of disease is described by patients through a shaded drawing and a description of the affected area in terms of the size of their hand, which is then assessed by a physician for percentage of affected BSA. The severity is assessed by the dryness of noninflammed skin and the characteristics of inflamed skin, through characteristics such as erythema, edema, crusts/oozing, excoriations, licheniifcation, bleeding, scaling, and flaking. Subjective items pruritus and sleep disturance are assessed using a visual analog scale. | Adults and children | The PO-SCORAD score defines three classes of severity: minor, moderate, and severe AD. | The PO-SCORAD score is a composite score that combines the assessments of the scale's three sections, ranging from 0 to 103. PO-SCORAD is defined by 0.2A + 3.5B + C, $A \in [0,100]$ corresponds to the extent, $B \in [0,18]$ to intensity signs, and $C \in [0,20]$ to subjective symptoms. A score between 0-25 is a minor case of AD; a score between 25-50 is a moderate case of AD; and a score above 50 is a severe case of AD. |
| POEM (Patient-Oriented Eczema Measure) | POEM is a patient-reported tool used to monitor atopic eczema severity, suitable for use in outpatient clinics, audit, research, and clinical trials. The criteria to dermine use on patients include the diagnosis of atopic dermatitis based on Hanifin-Rajka criteria; and the patient's ability to self-report frequency of atopic dermatitis symptoms. It is structured into a survey with seven questions regarding the frequency of pruritus, sleep disturbance, bleeding, oozing, cracking, flaking, and dryness over a one week period. | Adults and children | The POEM score defines five strata of severity: clear or almost clear; mild eczema; moderate eczema; severe eczema; and very severe eczema. | POEM incorporates seven symptoms into the final patient-oriented eczema measure using a 5-point scale of frequency of occurrence during the one-week period, with a maximum total score of 28. They are scored as clear or almost clear (0-2), mild eczema (3-7), moderate eczema (8-16), severe eczema (17-24), and very severe eczema (25-28). |
| EASI (Eczema Area and Severity Index) | EASI is a tool used to measure the extent and severity of atopic dermatitis, suitable for clinical practice and clinical trials. This scale requires a clinical diagnosis of atopic dermatitis and a full-body skin examiniation by a dermatologist or trained assessor to rate the severity and extent of lesions on four body regions (head and neck, trunk, upper limbs, and lower limbs). | Adults and children (multipliers differ slightly for ≤ 8 years old) | The EASI score ranges from 0-72. J.M. Hanifiin recommends stratifiction of clear or no eczema (0); almost clear (0.1-1.0); mild disease (1.1-7.0), moderate disease (7.1-21.0), severe disease (21.1-50.0), and very severe disease (<51). | EASI severity score multiplies the severity score by the area score. The severity score is the sum of the intensity scores for erythema (E), infiltration/papulation (I), excoriation (Ex), and lichenification (L). These intensities are assessed on a scale 0-3 indicating none (0), mild (1), moderate (2) and severe expression (3) of the clinical sign (half steps are allowed). The percent area of involvement of each of the four body regions is represented by a numeric value [0 (no eruption), 1 (<10%), 2 (10%-29%), 3 (30%-49%), 4 (50%-69%), 5 (70%-89%), 6 (90% -100%)]. The EASI can then be calculated according to the following formula: Upper limbs [Severity x Area × 0.2] + Lower Limbs [Severity x Area × 0.4] + Trunk [Severity x Area × 0.3] + Head/Neck [Severity × Area x 0.1] = EASI. The EASI score ranges from 0-72. J.M. Hanifiin recommends stratifiction of clear or no eczema (0); almost clear (0.1-1.0); mild disease (1.1-7.0), moderate disease (7.1-21.0), severe disease (21.1-50.0), and very severe disease (<51). |
| SCORAD (Scoring of Atopic Dermatitis) | SCORAD is a clinical scale used to assess the extent and severity of atopic dermatitis, suitable in clinical practice and clinical trials. It requires the diagnosis of atopic dermatitis based on clinical examination and assessment by trained clinicians. Administered by a dermatologist or trained assessor, six body regions (head and neck, upper limbs, lower limbs, anterior trunk, back, and genitals) are scored via a shaded drawing to determine extent. Intensity scores as are assessed for redness, swelling, oozing/crusting, scratch marks, lichenification, and dryness. Subjective symptoms of itch and sleepnlessness are patient-reported with a visual analog scale. | Adults and children. | The SCORAD score ranges from 0-103 and defines three classes of severity: minor, moderate, and severe AD. | The SCORAD index formula is A/5 + 7B/2 + C. In this formula A is defined as the extent (0-100), B is defined as the intensity (0-18) and C is defined as the subjective symptoms (0-20). A score between 0-25 is a minor case of AD; a score between 25-50 is a moderate case of AD; and a score above 50 is a severe case of AD. |
| I-DQoL (Infant's Dermatitis Quality of Life Index) | I-DQoL is a questionnaire completed by parents of children under the age of four years to assess the impact of atopic dermatitis on the quality of life in a one-week period. It is suitable for clinical practice and clinical trials. The questionnaire contains 10 questions on symptoms and difficulties with mood, sleep, play, family activities, mealtimes, treatments, dressing, and bathing. There is an additional question which is scored separately, asking for the parents' assessment of current dermatitis severity. | ≤ 4 years old | The I-DQoL score ranges from 0-30, and defines a dermatitis severity score and a life quality index. Dermatitis severity strata ranges from extremely severe (4), severe (3), average (2), fairly good (1), and none (0). The life quality index does not have formal classes. | 10 items scored 0-3. Score is summed up, max 30. There is also a subjective question scored 0-4. |

| | | | | |
|---|---|---|---|---|
| RECAP (Recap of Atopic Eczema) | RECAP is a patient-reported questionnaire that captures assessment of long-term control of eczema, intended for individuals with eczema of all ages in clinical trials. It is structured with seven quality of life questions regarding acceptability of eczema, itchy skin, sleep disturbance, quality of life in day-to-day activities, mood, and intensity of itch. There are two versions; a self-reported questionnaire for adults and older children with atopic dermatitis, and a caregiver-reported questionnaire for younger children with atopic dermatitis. | Adults and children | The RECAP score ranges from 0-28, defining how controlled the disease is. Bandings for interpretation of the scores has been reported as: completely controlled (0-1), mostly controlled (2-5), moderately controlled (6-11), a little controlled (12-19), and not at all controlled (20-28). | Each of the seven questions in RECAP carries equal weight and is scored from 0-4, witha total range of 0-28. |
| ADCT (Atopic Dermatitis Control Tool) | ADCT is a patient-reported outcome measure that captures atopic dermatitis control, as perceived by the patient. It is intended for use in clinical trials and routine clinical practice for adults and adolescents (12 years and older). The questionnaire is structured with six questions, spanning six concepts to measure atopic dermatitis control: overall severity of symtpoms, frequency of intense episodes of itcing, intensity of bother, frequency of sleep impact, intensity of daily activities impact, and intensity of mood or emotions impact. | ≥ 12 years old | The ADCT score ranges from 0-24, defining how controlled the disease is. A higher score indicates lower atopic dermatitis control, with a score of at least 7 points suggesting limited control. A change of 5 points is the threshold for meaningful within-person change. | Each of the six questions in ADCT carries equal weight and is scored from 0-4 points, with a total range of 0-24. |
| vIGA-AD (Validated Investigator Global Assessment for Atopic Dermatitis) | vIGA-AD is a standardized clinician-rated scale to assess the overall severity of AD lesions across all age groups, used in clinical practice and clinical trials. It is scored on a 5-point investigator global assessment scale based on AD lesions' erythema, induration/papulation, lichenification, and oozing/crusting, taking the extent of disease into account. | Adolescents, adults, and children | The vIGA-AD score ranges from 0-4, defining the morphological features of atopic dermatitis lesions. It is stratified by clear (0), almost clear (1), mild (2), moderate (3), and severe (4). | Lesions are assessed on a 5-point IGA scale of 0-4. |
| o-SCORAD (objective SCORAD) | Derived from SCORAD, o-SCORAD is a clinical scale used to assess the extent and severity of atopic dermatitis without including subjective symptoms, which have unpredictable impact on the end score. It requires the diagnosis of atopic dermatitis based on clinical examination and assessment by trained clinicians. Administered by a dermatologist or trained assessor, six body regions (head and neck, upper limbs, lower limbs, anterior trunk, back, and genitals) are scored via a shaded drawing to determine extent. Intensity scores as are assessed for redness, swelling, oozing/crusting, scratch marks, lichenification, and dryness. | Adults and children. | The o-SCORAD score ranges from 0-93 and defines three classes of severity: minor, moderate, and severe AD. | The o-SCORAD index formula is $A/5 + 7B/2$. In this formula A is defined as the extent (0-100) and B is defined as the intensity (0-18). Bonus points are added for disfiguring lesions or function-limiting lesions, up to 10 points. A score between 0-25 is a minor case of AD; a score between 25-50 is a moderate case of AD; and a score above 50 is a severe case of AD. |
| PSAAD (Pruritus and Symptoms Assessment for Atopic Dermatitis) | PSAAD is a patient-reported outcome tool that assesses daily AD symptoms during the course of therapy, aiming to establish content validity and psychometric properties to support product-labelling claims. The scale requires patients to be aged ≥ 12 years and have a clinical diagnosis of moderate-to-severe AD.The structure of the questionnaire contains 11 items, assessing itch, dryness, pain, flaking, cracking, bumps, redness, discolouration, bleeding, fluid, and swelling over 24 hours. | ≥ 12 years old | The PSAAD score ranges from 0-10, with no formal stratification. | Each item of the PSAAD assesses the severity of a single symptom on an 11-point NRS, ranging from 0 (none) to 10 (extreme), and contributes equally to the PSAAD total score. The PSAAD total score is calculated as the average of the responses to each of the 11 items, for a PSAAD total score range of 0 (none) to 10 (extreme). |
| IGAxBSA (Product of Investigator Global Assessment and Body Surface Area) | IGAxBSA is the multiplied product of vIGA-AD and BSA, capturing a clinician's assessment of AD lesion severity and extent of disease through percentage of lesioned body surface area. This scale is aimed towards children for use in clinical settings. | ≤ 17 years old | The IGAxBSA score ranges from 0-400, with a suggested severity strata of mild (0-30), moderate (30.1-130), and severe, (130.1-400). | The end score is the product of the vIGA-AD and BSA. The vIGA-AD is scored on a scale of 0-4, and then multiplied by the BSA. |
| Rajka–Langeland severity score | The Rajka-Langeland score is a 4-tem scale that aims to grade the severity of atopic dermatitis through extent of disease (localized or widespread), symptom intensity (itch, dryness, redness, swelling, crusting), and frequency/chronicity of flare-ups. It is suitable for use in clinical practice and clinical trials. | Adults, children, and infants | The Rajka-Langeland score ranges from 3-9, with strata of mild (3-4), moderate (4.5-7.5), and severe (8-9). | Rajka–Langeland scores (range 3–9) were calculated by summing the scores for remission, itch intensity and BSA (1, < 9%; 2, 10–35%; 3, ≥ 36%). |

| | | | | |
|---|---|---|---|---|
| Atopic Dermatitis Screening and Evaluation Questionnaire (ADSEQ) | ADSEQ is a patient-reported questionnaire designed to validate self-reported atopic dermatitis diagnosis, assess self-reported AD severity, and measure itch serverity, distinguishing it from other skin conditions. It is structured with 13 questions accompanied by representative images of atopic dermatitis on five different body locations. The questions involve clinical diagnosis, history of atopic disease, dry skin, itchy skin, itch frequency and intensity, rash locations, and lesion morphology. | Adults | ADSEQ provides a diagnosis of atopic dermatitis. | No numerical values included. |
| Childhood Eczema Questionnaire (CEQ) | CEQ is a parent-reported questionnaire for diagnosing atopic dermatitis in children up to two years of age. It contains questions characteristic to atopic dermatitis regarding intermittent red rashes, itching, location of lesions, and dry skin. It is suitable for use in clinical practice and clinical trials. | 1 month to 2 years old | CEQ provides a diagnosis of atopic dermatitis. | No numerical values included. |
| Patient-Reported Outcomes Measurement Information System (PROMIS) Itch Questionnaire (PIQ) | PROMIS PIQ is a set of item banks designed to improve assessment of health-related quality of life in itch, covering general concenrs, mood and sleep, clothing and physical activity, and scratching behaviour. It is administered individually or in combination within clinical practice and clinical trials across adult, pediatric, and proxy populations. | Adults, children, and proxy | PROMIS PIQs assess various outcomes regarding pruritus-related quality of life. | Scores are reported on a T-score metric and range from 32.7-79.1 for item bank 1, 26.9-77.4 for bank 2, 32.6-77.1 for bank 3, and 32.6-72.7 for bank 4. A T-score of 50 is the average score for people in the United States general population who recently experienced chronic itch for any reason. |
| Atopic Dermatitis Severity Index (ADSI) | ADSI is a tool consisting of the sum of scores for pruritus, erythema, exudation, excoriation, and lichenification. The tool focuses on arm lesions and assesses the severity of the disease. | Not specified | ADSI scores are used over time to determine change from baseline lesion severity. | Each symptom is scored on a 4-point rage from 0-3 and summed for the final ADSI score. |
| Childhood Atopic Dermatitis Impact Scale (CADIS) | CADIS is a parent-reported instrument that measures the effects of atopic dermatitis on the quality of life of young children under 6 years of age and their families, with suitability for clinical research. The questionnaire includes 45 questions about the child's eczema during a 4-week period, itch intensity, sleep loss, overall skin condition, and quality of life for both the patients and guardians. | Parents and patients ≤ 6 years old | Establishing the severity of the impact of atopic dermatitis on quality of life, CADIS scores can be reported overall or within their individual domains of child symptoms, child activity limitations and behaviour, family and social function, parent sleep, and parent emotions. | Standardized response choices are scored on a 5-point scale ranging from 0-4. Items about itch and sleep loss were rated on 10-point visual analog scales. Questions are weighted equally and summed for a total CADIS and individual domain score. |
| Quality of Life Index for Atopic Dermatitis (QoLIAD) | QoLIAD is a patient-reported outcome measure assessing the quality of life of adult and adolescent patients with atopic dermatitis, suitable for clinical practice and clinical trials. It contains 25 items with a dichotomous response system and questions centering around the need for mental and emotional stimulation; physical and emotional stability; security; sharing and belonging; esteem; and personal development and fulfilment. | ≤ 16 years old | QoLIAD scores are summed by affirmative responses to the questions, with a higher score indicating a greater negative impact of atopic dermatitis on the patient's quality of life. | Each question is scored dichotomously and summed for the final QoLIAD score, ranging from 0-25. |
| Parents' Index of Quality of life in Atopic Dermatitis (PIQoL-AD) | PIQoL-AD is a quality of life instrument specific to aprents of children with atopic dermatitis, assessing the impact of the disease and its treatment on the family. It is suitable for clinical practice and clinical trials. It contains contains 28 items with a dichotomous response system and questions centering around the need for child to have a safe and successful life; rest and relaxation; self-respect; independence; personal space and time; and control. | Parents of children ≤ 8 years old with atopic dermatitis | PO-QoLIAD scores are summed by affirmative responses to the questions, with a higher score indicating a greater negative impact of atopic dermatitis on the family's quality of life. | Each question is scored dichotomously and summed for the final PO-QoLIAD score, ranging from 0-28. |
| Dermatitis Family Impact (DFI) | DFI is a quality of life measure used in clinical practice that assesses the impact on quality of life of atopic dermatitis in infants and their families. The questionnaire is structured with 10 questions on a 4-point scale ranging from 0-3, with a maximum score of 30. Questions involve housework, food preparation and feeeding, sleep, family leisure activity, shopping, expenditure, tiredness, emotional distress, relationships, and impact of helping with treatment on the main carer's life in a one-week period. | Caregivers of children with AD | DFI scores are summed by affirmative responses to the questions, with a higher score indicating a greater negative impact of atopic dermatitis on the family's quality of life. | Each question is scored dichotomously and summed for the final DFI score, randinging from 0-30. |
